# Pseudo-Label Assisted Nnu-Net (PLAn) Enables Automatic Segmentation of 7T MRI From a Single Acquisition

**DOI:** 10.1101/2022.12.22.22283866

**Authors:** Henry Dieckhaus, Corinne Donnay, María Inés Gaitán, Erin S. Beck, Andrew Mullins, Daniel S. Reich, Govind Nair

## Abstract

**Introduction:** Automatic whole brain and lesion segmentation at 7T presents challenges, primarily from bias fields and susceptibility artifacts. Recent advances in segmentation methods, namely using atlas-free and multi-contrast (for example, using T_1_-weighted, T_2_-weighted, fluid attenuated inversion recovery or FLAIR images) can enhance segmentation performance, however perfect registration at high fields remain a challenge primarily from distortion effects. We sought to use deep-learning algorithms (D/L) to do both skull stripping and whole brain segmentation on multiple imaging contrasts generated in a single Magnetization Prepared 2 Rapid Acquisition Gradient Echoes (MP2RAGE) acquisition on participants clinically diagnosed with multiple sclerosis (MS). The segmentation results were compared to that from 3T images acquired on the same participants, and with commonly available software packages. Finally, we explored ways to boost the performance of the D/L by using pseudo-labels generated from trainings on the 3T data (transfer learning).

**Methods:** 3T and 7T MRI acquired within 9 months of each other, from 25 study participants clinically diagnosed with multiple sclerosis (mean age 51, SD 16 years, 18 women), were retrospectively analyzed with commonly used software packages (such as FreeSurfer), Classification using Derivative-based Features (C-DEF), nnU-net (“no-new-Net” version of U-Net algorithm), and a novel 3T-to-7T transfer learning method, Pseudo-Label Assisted nnU-Net (PLAn). These segmentation results were then rated visually by trained experts and quantitatively in comparison with 3T label masks.

**Results:** Of the previously published methods considered, nnU-Net produced the best skull stripping at 7T in both the qualitative and quantitative ratings followed by C-DEF 7T and FreeSurfer 7T. A similar trend was observed for tissue segmentation, as nnU-Net was again the best method at 7T for all tissue classes. Dice Similarity Coefficient (DSC) from lesions segmented with nnU-Net were 1.5 times higher than from FreeSurfer at 7T. Relative to analysis with C-DEF segmentation on 3T scans, nnU-Net 7T had lower lesion volumes, with a correlation slope of just 0.68. PLAn 7T produced equivalent results to nnU-Net 7T in terms of skull stripping and most tissue classes, but it boosted lesion sensitivity by 15% relative to 3T, increasing the correlation slope to 0.90. This resulted in significantly better lesion segmentations as measured by expert rating (4% increase) and Dice coefficient (6% increase).

**Conclusion:** Deep learning methods can produce fast and reliable whole brain segmentations, including skull stripping and lesion detection, using data from a single 7T MRI sequence. While nnU-Net segmentations at 7T are superior to the other methods considered, the limited availability of labeled 7T data makes transfer learning an attractive option. In this case, pre-training a nnU-Net model using readily obtained 3T pseudo-labels was shown to boost lesion detection capabilities at 7T. This approach, which we call PLAn, is robust and readily adaptable due to its use of a single commonly gathered MRI sequence.

## Introduction

Volumetric segmentation of brain magnetic resonance images (MRI) is now a commonly used postprocessing step enabling noninvasive quantitative analysis of disease progression in various neurological and neuropsychiatric diseases such as multiple sclerosis (MS),(1) human immunodeficiency virus,(2) Alzheimer’s disease,(3) and depression.(4) Since manual annotation of 3D MRI data is extremely tedious and time-consuming, automated segmentation methods are of great interest for both clinical and research applications. There is a dearth of brain segmentation algorithms from 7T images compared to 3T and lower fields, especially ones that can efficiently segment tissue classes in the whole brain.

Higher magnetic fields (7T, compared to 3T) offer higher signal-to-noise ratio, enabling submillimeter resolution, which in turn allows more sensitive analyses than were previously possible.(5) For instance, cortical lesions are associated with cognitive decline in MS but are not detected as reliably at 3T as they are at 7T.(6) The greater structural detail enabled by 7T imaging also allows more nuanced stratification of pathology such as MS lesions or glioblastomas.(7) However, higher magnetic fields also present particular challenges for postprocessing as a result of more pronounced radiofrequency field nonuniformities as well as more susceptibility artifacts (8) and larger spatial distortion near air-tissue interfaces. These complicate coregistration, and any misregistrations can adversely affect multi-contrast segmentation efforts.(9) In addition, fluid-attenuated inversion recovery (FLAIR) images, typically extremely useful in detecting lesions at 3T and lower field strengths, are less useful at 7T due to generally less lesion conspicuity and stronger bias fields from multiple, strong RF pulses. (10, 11) As a result, multi-contrast segmentation methods developed for lower field strengths, such as FreeSurfer and Classification using Derivative-based Features (C-DEF), may be degraded in their effectiveness when applied to 7T data.(12) Sequences such as multi-echo GRE and Magnetization Prepared 2 Rapid Acquisition Gradient Echoes (MP2RAGE) offer the opportunity to obtain multiple imaging contrast in the same acquisition, which not only can have similar distortions but also alleviate the need for registration between images.(13) In addition, incorrect skull stripping, especially affecting 7T scans, can also contribute to inaccurate brain segmentation. The cerebrospinal fluid (CSF) class, a surrogate marker of whole brain atrophy, is often the most affected by over- or under-stripping errors. Furthermore, tissue outside the brain may be erroneously classified as part of the brain if included in the brain mask due to under-stripping. It is therefore important to incorporate accurate skull stripping techniques into brain segmentation algorithms to improve its accuracy.

Deep learning, especially convolutional neural networks (CNNs) derived from the U-Net framework, have become the state-of-the-art for brain MRI segmentation in recent years.(14) These methods rely on layers of automatically optimized filters and nonlinear activations that enable learning sophisticated image features from an annotated training dataset, which are then applied to predict on unseen data. In particular, the nnU-Net method introduced by Isensee *et al*.(15) has become the *de facto* baseline segmentation method for a wide range of medical segmentation tasks. A few prior studies have attempted to apply CNNs to segment high-field MRI data. Custom CNNs have been used for cortical lesion segmentation (16) and multiclass whole-brain segmentation (12) on 7T data. For this study, we hypothesized that the capabilities of nnU-Net, perhaps boosted by domain-specific adaptation, may reduce the dependence on auxiliary image contrasts or *a priori* information by relying instead on contextual information extracted from the training dataset.

This manuscript presents a deep-learning method for atlas-free, automatic, whole-brain segmentation of 7T MRI, integrating the interdependent tasks of skull stripping and lesion segmentation. The novel method, Pseudo-Label Assisted nnU-Net (PLAn), wherein a nnU-Net deep learning model is pre-trained with readily obtained pseudo-label data derived from scans at a lower field strength (3T), then fine-tuned with limited 7T expert-drawn labels (transfer learning) in order to optimize lesion segmentation performance. The performance of this method was compared against manually drawn masks and other commonly available methods on 3T and 7T data acquired within 9 months, in patients clinically diagnosed with MS.

## Materials and methods

### Image Acquisition

The study protocol was approved by the institutional review board of the National Institutes of Neurological Disorders and Stroke (NCT: NCT00001248), and all participants provided informed written consent. MRI scans used in this study were from 3T and 7T (Siemens, Malvern, PA) performed within 9 months on participants in our natural history of MS study, who did not show any disease progression between the scans, and in whom no new lesions could be detected between the two scans based on radiological reads. 3T images, acquired on a Skyra system with a 32-channel head coil, included T_1_-weighted (T_1_w) images (MP2RAGE, TR/TE/TI_1_/TI_2_ = 5000/2.26/700/2500 ms, flip angle = 4.5°, 1-mm isotropic resolution), 3D FLAIR (TR/TE/TI = 5000/393/1800 ms, 1-mm isotropic resolution), and PD/T_2_ (2D FSE, TR/TE_1_/TE_2_ = 3630/9.6/96 ms, resolution = 0.7×0.7×3 mm). 7T images were acquired on a Magnetom system equipped with a 1-channel transmit/32-channel receive coil and included T_1_w images and T_1_ maps (MP2RAGE, TR/TE/TI_1_/TI_2_ = 4000/4.6/350/1350 ms, flip angle = 4.5°, 0.7-mm isotropic resolution).

### Other methods for performance comparison

Schematic diagrams of the workflow is shown in Supplementary Figure 1. Methods that were evaluated for skull stripping included 3dSkullStrip tool (*shrink_fact_bot_lim* = 0.7, AFNI toolkit)(17) on the 3T (AFNI 3T) and 7T (AFNI 7T) MP2RAGE second inversion images. Skull-strip outputs from deep learning methods were evaluated against these.

Segmentation methods initially evaluated included not only FreeSurfer and C-DEF on 3T and 7T images but also nnU-Net applied to 7T images. The uniformized-denoised T_1_w, inversion 1, and inversion 2 images from MP2RAGE scan, along with FLAIR from 3T were inputs for C-DEF (C-DEF 3T) while T_1_ map, inversion 1, and inversion 2 images from MP2RAGE scan from 7T served as inputs for the nnUNET (nUNET 7T) and C-DEF (C-DEF 7T). Similarly, the T_1_w and FLAIR scans from 3T were used for FreeSurfer segmentation (FreeSurfer 3T), while just the T_1_w scans from 7T were used as input for FreeSurfer 7T after down-sampling 7T T_1_w MP2RAGE scans to 1.0 mm^3^, processing by *recon-all*, then up-sampling back to 0.7 mm^3^ space. MRI images were preprocessed prior to modeling with C-DEF; preprocessing included coregistration and bias-field correction using a sliding percentile filter.(18) This was followed by 5-fold cross-validation on 5 manually annotated training scans and ensembled inference using majority voting. All 7T scans were corrected for bias-field inhomogeneity using ANTs N4 bias correction (19) prior to modeling. Due to memory constraints at higher resolutions, FreeSurfer segmentations on 7T (FreeSurfer 7T) were obtained by FreeSurfer automatic subcortical segmentation outputs were converted into NIfTI format, then mapped from various anatomical labels to one of four tissue classes: CSF, grey matter (GM), WM, and lesions. All C-DEF and FreeSurfer processing was implemented on a computing cluster with 64x Intel^®^ Xeon^®^ CPUs with 256 GB RAM running Centos 7.7.

To create 7T training labels for deep learning methods, manually drawn lesion masks and skull stripping masks were added to FreeSurfer 7T grey and WM masks, which were then heavily edited for errors (CD, HD) and validated by trained and experienced neurologists (E.S.B. or M.I.G.). C-DEF 7T segmentations were obtained by the same procedure as previously described for C-DEF 3T, except that the percentile filter bias correction was omitted in favor of the N4 bias correction and per-image z-score normalization was applied during preprocessing. Finally, nnU-Net 7T segmentations were obtained using the publicly available nnU-Net package (15) after running the full cross-validation, model selection, and ensembled inference routine as described in the nnU-Net documentation (https://github.com/MIC-DKFZ/nnUNet). The only significant modification made to nnU-Net 7T was to disable largest-connected-component postprocessing, which was found to not be helpful for this task. All nnU-Net models were implemented on a computing cluster with NVIDIA^®^ v100-SXM2 GPUs each with 32 GM VRAM and 24 Intel^®^ Xeon^®^ CPUs each with 64 GB RAM.

### Pseudo-Label Assisted nnU-Net (PLAn) 7T

To obtain the 3T pseudo-labels, results were obtained from C-DEF 3T as previously described, then up-sampled to 0.7 mm^3^ using cubic interpolation for input images and nearest neighbor interpolation for labels. Both the training set (*n* = 5) and testing set (*n* = 20) were split into 5 folds for cross-validation. Pseudo-labels and 3T MP2RAGE scans were used for pre-training of an nnU-Net model using the 3D full-resolution U-Net configuration and default settings. Transfer learning was achieved by loading the pre-trained model weights and preprocessing settings (except for the final softmax layer), then fine-tuning the network with the manually edited 7T labels and 7T MP2RAGE scans. Fine-tuning was conducted for 125 epochs (default = 1000) with an initial learning rate of 1 × 10^−4^ (default = 1 × 10^−2^), as preliminary analysis found it to be more efficient and accurate (learning rate optimization results are shown in Supplementary Figure 2). No other settings were altered, and no model layers were frozen during fine-tuning. The fine-tuned model was then applied to the 7T MP2RAGE scans of the remaining unseen participants to obtain cross-validation results.

### Qualitative Evaluation

Segmentation outputs were randomly scrambled, and the method to generate each segmentation method was hidden prior to qualitative evaluation. An experienced neurologist (M.I.G.) rated the quality of segmentation for each tissue class, as well as for skull stripping, for each participant scan. A subjective rating scale from 1 to 5 was used with the best segmentation being ranked as a 5. For example, if all the lesions in a participant was correctly segmented matching the exact shape of the lesions, a score of 5 would be assigned to the method for lesions segmentation from that participant. Similarly, if most of a tissue class was wrongly assigned at segmentation, a score of 1 would be assigned for that tissue class and participant. The mean tissue score and 95% confidence interval from all participants for each method was then calculated and compared.

### Quantitative Evaluation and Statistical Analysis

Dice similarity coefficient (DSC) was calculated for each tissue class as described in Zou et al. with various reference images as stated for each experiment,(20) while tissue volumes were calculated using the *fslstats* utility from FSL.(21) Statistical analysis was performed using PRISM^®^ version 9.3.1. After checking the normality assumption using the Shapiro-Wilk test, repeated-measures ANOVA was used to determine significant changes in mean metrics between segmentation methods, using Dunnett’s method to correct for multiple comparisons. A corrected P-value of 0.05 or lower was considered statistically significant. Correlation of tissue volumes between methods was calculated using Pearson’s correlation coefficient, while Bland-Altman analysis was used to determine mean bias and 95% limits of agreement between methods. Unless otherwise noted, all statistics are given in terms of their mean value and 95% confidence intervals (CI). All data reported are in the form of mean ± standard deviation, unless mentioned otherwise.

### Data availability

The nnU-Net package is publicly available on GitHub (https://github.com/MICDKFZ/nnUNet). The data and code used here may be obtained with Material Transfer Agreements, subject to our institutes policy on data sharing.

## Results

### Demographic and Clinical Data

MRI from 25 participants of an IRB-approved natural history of MS study (age: 51 ± 16 years, sex: 18 women / 7 men, MS phenotype: 15 relapsing-remitting, 8 secondary progressive, 1 primary progressive, 1 other neuroinflammatory disease with MS-mimicking brain lesions), meeting inclusion criteria as mentioned in the methods section, were used in this study. All participants provided signed, informed consent to participate in the study. The training/validation (*n* = 5) and testing (*n* = 20) data sets were acquired on 3T and 7T within a median 36 days (range: 1 – 196 days). Participants were selected from the full study to ensure that no new lesions were visible by MRI between the two scans.

### Skull stripping

The nnU-Net 7T and PLAn 7T methods both produced excellent skull-stripping results throughout the brain (Fig. 1A–D). When compared volumetrically to the AFNI 3T reference method by Bland-Altman analysis (Fig. 1E), both methods had very low mean TIV biases, with 0.88% (CI: -3.8, 5.6) for nnU-Net 7T and 0.89% (CI: -3.7, 5.5) for PLAn 7T. Qualitative evaluation revealed that deep learning methods offered slight improvements over AFNI at 7T, particularly in dorsal (Fig. 1A), retro-orbital (Fig. 1B), and cerebellum (Fig. 1C) regions. Expert ratings (with 1 being poor and 5 being accurate segmentation, Table 1) confirmed these observations, with mean skull stripping scores of 3.84 (CI: 3.66, 4.02) for nnU-Net 7T and 3.80 (CI: 3.61, 3.99) for PLAn 7T, respectively, compared to just 3.11 (CI: 2.83, 3.80) for AFNI 7T skull stripping. Indeed, Bland-Altman analysis comparing AFNI 7T to AFNI 3T (Fig. 1E) produced a mean TIV bias of -3.5% (CI: -8.2, 1.1), which is several times greater in magnitude than that from either deep learning method. While AFNI 7T and AFNI 3T produced similar skull stripping boundaries in some brain regions, such as dorsal regions (Fig. 1A), other regions such as cerebellum showed significant degradation at 7T compared to 3T (Fig. 1C). FreeSurfer 7T, on the other hand, was prone to overestimation of brain volumes throughout the entire brain, including extracranial areas abutting the anterior temporal lobe (Fig. 1D) and cerebellum (Fig. 1C). This resulted in a mean TIV bias of 10% (CI: 3, 19) when compared to AFNI 3T (Fig. 1E). These errors were evidently more detrimental than those displayed by AFNI 7T, as expert ratings of skull stripping quality produced a mean score of just 1.63 (CI: 1.30, 1.96) for FreeSurfer 7T (Table 1).

**Figure 1.**
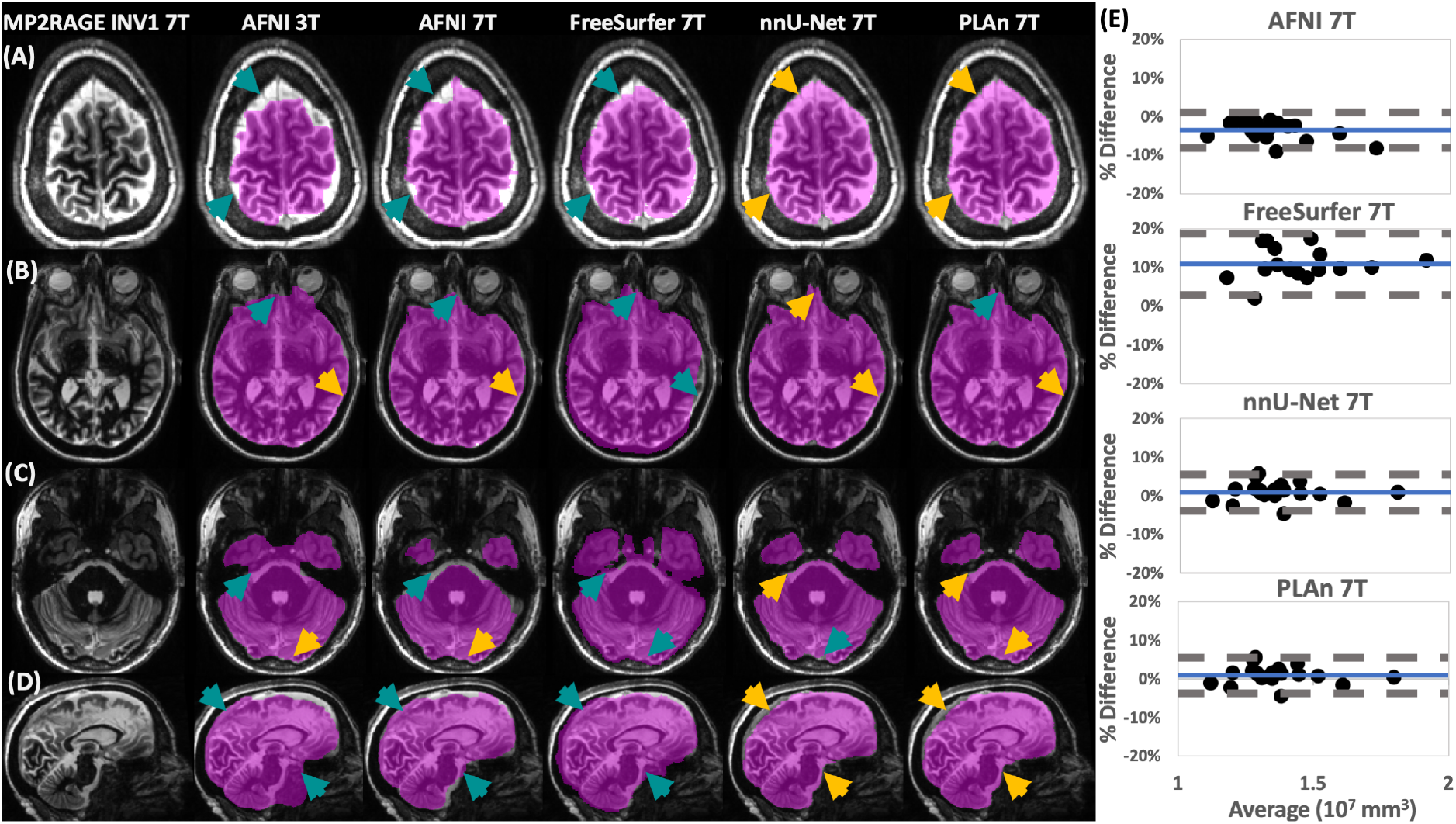
Qualitative and quantitative assessment shows nnU-Net and PLAn methods give superior skull stripping results at 7T. At left, representative **(A–C)** axial and **(D)** sagittal image slices selected from various participants from the inference set show the output of skull stripping methods (magenta overlay indicates regions identified as brain, method and field strength indicated at the top) in various brain regions overlaid on 7T MP2RAGE INV1 image. Gold arrows indicate good skull stripping features in areas where other methods made errors (teal arrows). At right, **(E)** Bland-Altman plots of total intracranial volume (TIV) calculated from 7T methods in A–D compared to AFNI 3T, which was used as a reference (% Difference = 100 x [Method – C-DEF 3T] / Average). Mean bias (solid blue) and 95% limits of agreement of the dataset (dashed grey) are indicated.

**Table 1:**
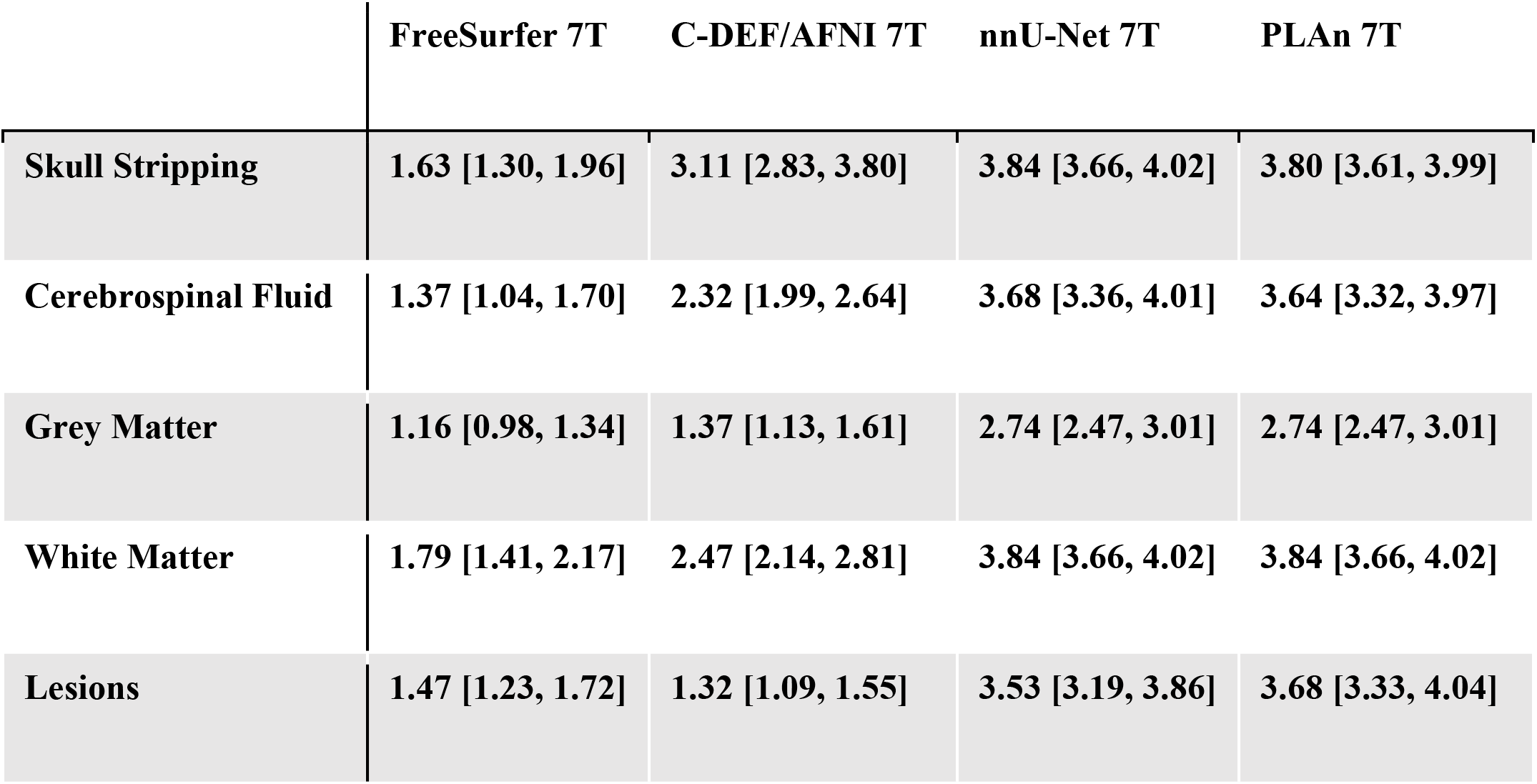
Blinded expert ratings (mean with 95% CI) for each 7T method for skull stripping and tissue segmentation. Rating scale is from 1 being poor and 5 being accurate segmentation for each class.

### Comparison of tissue segmentation methods (except PLAn)

The nnU-Net 7T significantly outperformed both C-DEF 7T and FreeSurfer 7T for all tissue segmentation classes (Fig. 2). It successfully captured detailed cortical boundaries (Fig. 2A– B) as well as cerebellum and brainstem borders (Fig. 2D). CSF volume correlation plots between nnU-Net 7T and C-DEF 3T (Fig. 3A) produced a strong correlation coefficient (*r* = 0.92, ([CI: 0.91, 0.97], *P* < 0.001, *slope* = 1.2). This is consistent with prior observations that C-DEF 3T under-strips the skull in dorsal regions, leading to slightly lower CSF volumes than expected.

**Figure 2.**
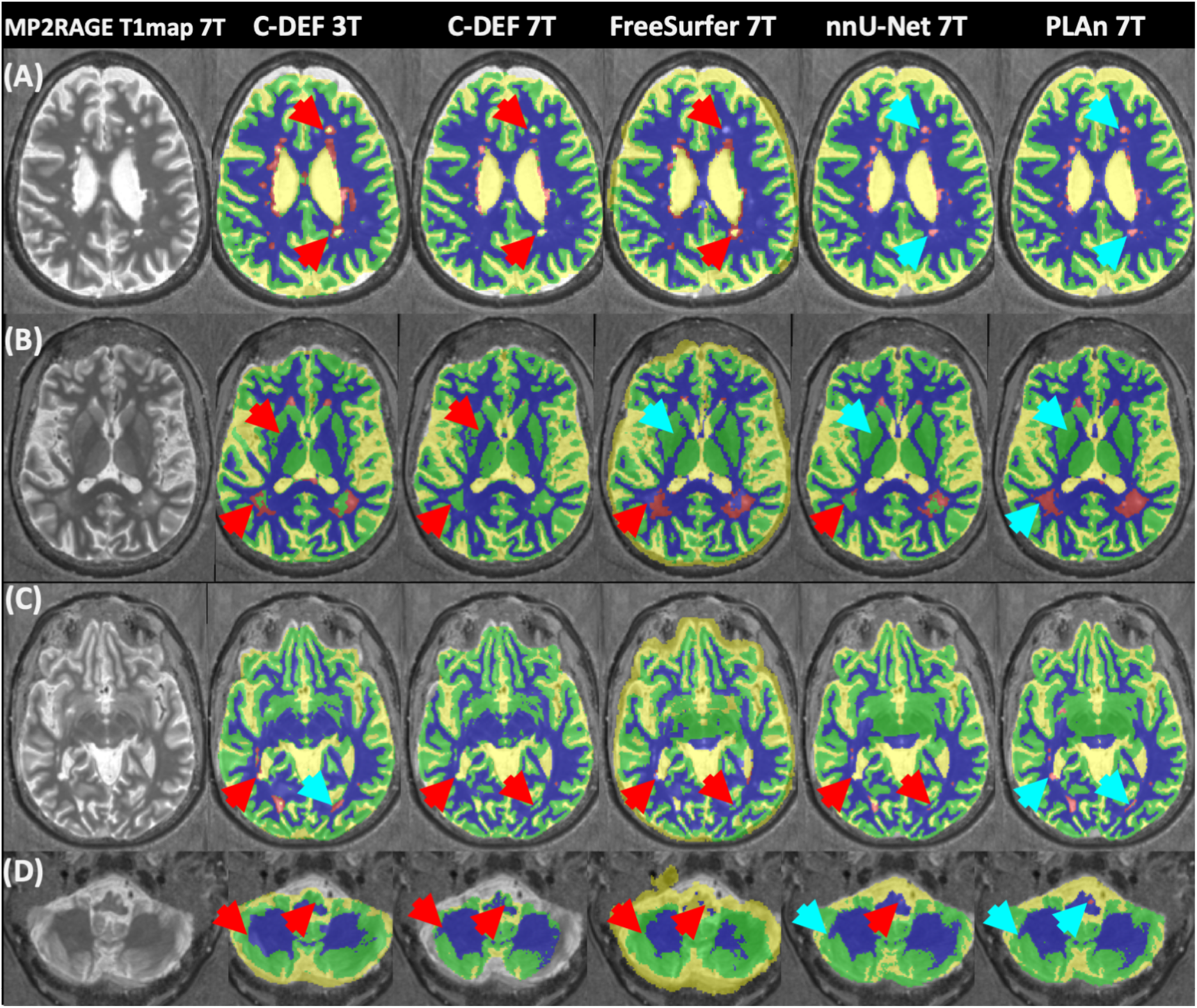
Qualitative assessment shows that PLAn 7T produces the best overall segmentations at 7T. Representative axial image slices selected from various participants in the inference set show different tissue segmentation methods (method and field strength indicated at the top) from various brain regions overlaid on 7T MP2RAGE T_1_ map image. Blue arrows indicate good segmentation features, red arrows indicate errors. For the segmentation overlays, blue indicates WM, yellow CSF, green GM, and magenta lesions.

**Figure 3.**
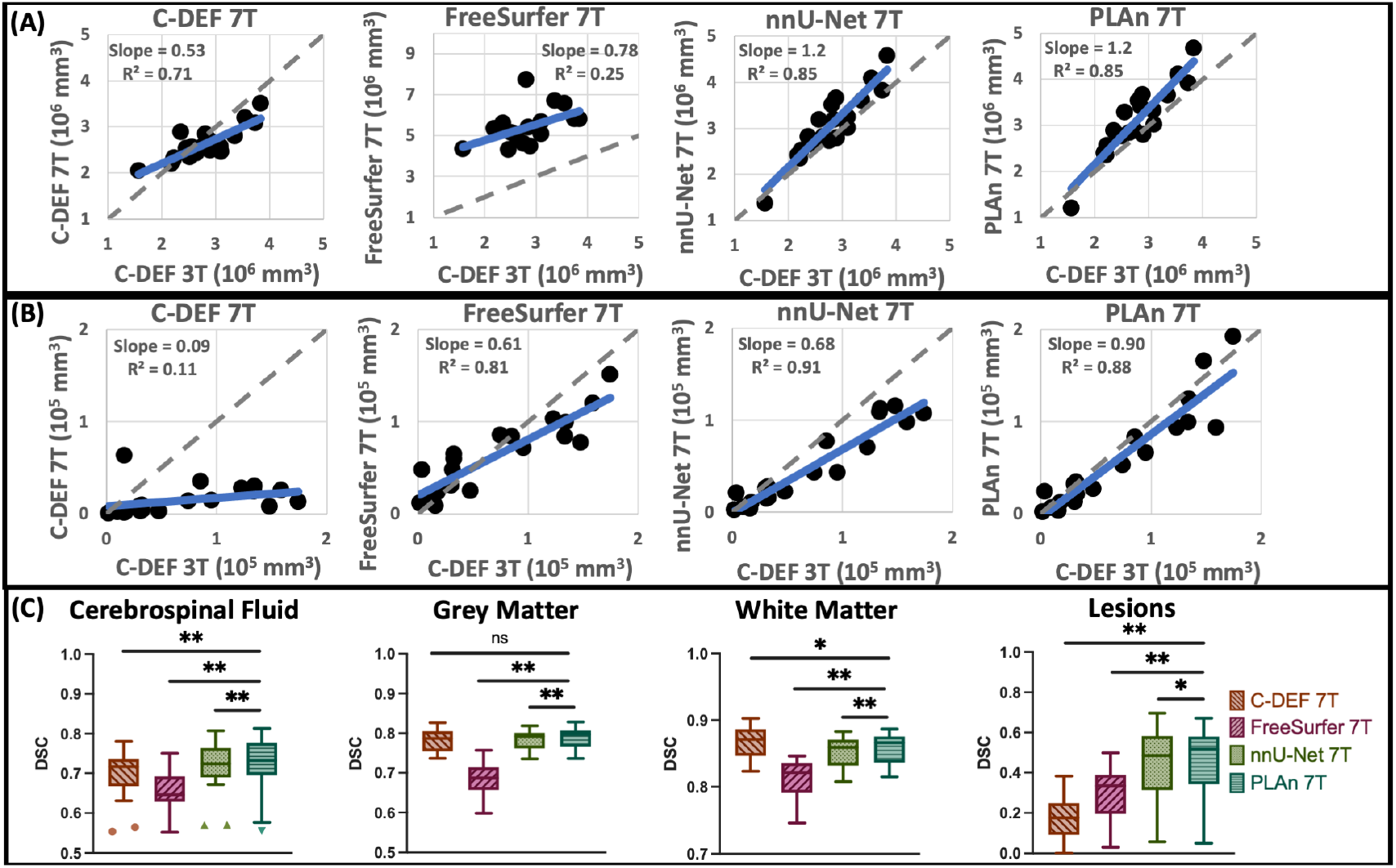
Quantitative assessment shows that PLAn 7T produces the best overall segmentations at 7T. Correlation plots of **(A)** CSF volume and **(B)** lesion volume of various segmentation methods (as indicated on top) vs C-DEF 3T reference with identity (dashed grey) and regression (solid blue) lines. **(C)** Dice similarity coefficient (DSC) plots of each tissue class vs C-DEF 3T reference; asterisks mark significant mean difference compared to PLAn 7T (* p < 0.05, ** p < 0.01).

FreeSurfer 7T produced degraded temporal lobe and cerebellum segmentations, with sulcal GM being mislabeled as CSF throughout the ventral regions (Fig. 2C–D). This resulted in highly inflated and unreliable CSF volumes, with a mean bias of 63% (CI: 30, 96) along with a comparatively low correlation coefficient (*r* = 0.50 ([CI: 0.02, 0.79], *P* > 0.05, *slope* = 0.78) when compared to C-DEF 3T. C-DEF 7T had much better CSF segmentations due to good cortical boundaries (Fig. 2A–B), although it still presented a modest mean CSF volume bias of -4.7% (CI: -29, 20) when compared to C-DEF 3T. While this bias did have a significant effect, resulting in a mean CSF DSC of 0.70 (CI: 0.67, 0.73), compared to 0.72 (CI: 0.69, 0.75) for nnU-Net 7T, it was less detrimental than the errors in FreeSurfer 7T (mean DSC: 0.65 [CI: 0.63, 0.68]), with C-DEF 3T as reference. Expert ratings confirmed that nnU-Net was the best 7T method for CSF segmentation, with a mean rating of 3.68 (CI: 3.36, 4.01), compared to 1.37 (CI: 1.04, 1.07) for FreeSurfer 7T and 2.32 (CI: 1.99, 2.64) for C-DEF 7T (Table 1). The nnU-Net also matched or exceeded FreeSurfer 7T in distinguishing deep GM structures such as thalamus and globus pallidus, which C-DEF often missed (Fig. 2B). As a result, nnU-Net 7T had substantially better expert ratings than C-DEF 7T or FreeSurfer 7T for both GM and WM (Table 1).

Correlation plots (Fig. 3 A, B) show a strong correlation between nnU-Net 7T and C-DEF 3T in CSF and lesion volumes (*r* = 0.95 [CI: 0.89, 0.98], *P* < 0.001, slope = 0.68). C-DEF 7T, on the other hand, failed to detect most lesions throughout the brain (Fig. 2A–D), which resulted in very low lesion volumes (Fig. 3B) and a correlation of just *r* = 0.33 ([CI -0.13, 0.68], *P* > 0.05, slope = 0.09). Meanwhile, FreeSurfer 7T demonstrated significant lesion sensitivity, with a moderately strong lesion volume correlation of *r* = 0.90 ([CI: 0.75, 0.96], *P* < 0.001, slope = 0.61), both of which were significant increases compared to C-DEF 7T (Fig. 3B). However, qualitative inspection revealed these lesion segmentations to be frequently inaccurate (Fig. 2A– C). It was noted that lesion volumes of participants with low lesion loads were inflated, whereas those with high lesion loads were deflated (Fig. 3B). Along these lines, significant increase in mean DSC vs the other 7T methods were observed when scored against C-DEF 3T, especially in CSF and lesion class (Fig. 3C). This accuracy increase was most evident in small-to-medium punctate lesions (Fig. 2A, 2C). Both lesion DSC (mean: 0.30 [CI: 0.23, 0.36]) and expert ratings (mean: 1.47 [CI: 1.23, 1.72]) in FreeSurfer 7T were only slightly better than that of CDEF 7T (mean DSC: 0.17 [CI: 0.12, 0.22]; mean rating: 1.32 [CI: 1.09, 1.55]), despite far greater lesion volumes.

### PLAn 7T improves lesion segmentation over nnU-Net 7T

Despite its substantial advantages over other 7T methods, nnU-Net 7T still produced noticeable deficiencies in lesion detection, such as those seen in Fig. 2B or C. To address these deficiencies, we implemented the PLAn 7T method (workflow described in Supplementary Fig. 1). For the fine-tuning step, cross-validation parameter sweeps were conducted for total epochs (not shown) and initial learning rate (Supplementary Fig. 2), resulting in an optimal configuration of 125 epochs and an initial learning rate of 1 × 10^−4^. Mean DSC values calculated against CDEF 3T indicated that PLAn 7T outperformed nnU-Net 7T in terms of tissue segmentation (Fig. 3C). For CSF (1%), GM (0.6%), and WM (0.5%), this effect was relatively small but statistically significant (*P* < 0.001), while expert ratings (Table 1) found either no (GM, WM) or slight (CSF, -1%) differences between nnU-Net 7T and PLAn 7T for these tissue classes. Correlation plots of CSF volumes vs C-DEF 3T (Fig. 3A) also appeared virtually identical between the two methods.

Importantly, PLAn 7T produced significant improvement in lesion sensitivity compared to nnU-Net 7T (full slices in Fig. 2, magnified details in Fig. 4). This boost was consistent across various lesion types, including diffuse WM hyperintensities that nnU-Net 7T was often unable to distinguish from GM or WM (Fig. 2B, Fig. 4B–C), as well as periventricular lesions, which were often misclassified as CSF (Fig. 2C, Fig. 4A, 4C). There was also a noticeable improvement in lesion detection using PLAn compared to nnU-Net in inferior brain regions, including the temporal lobe (Fig. 2C), putamen (Fig 2C), and brainstem (Fig 2D). Lesion volume correlation plots (Fig. 3B) found that while the correlation coefficients of the two methods were very similar (*r* = 0.95, [CI: 0.89, 0.98], *P* < 0.001 for nnU-Net 7T, compared to *r* = 0.94 [CI: 0.85, 0.98], *P* < 0.001 for PLAn 7T), PLAn 7T gave a consistent boost to lesion volumes (mean bias: 15% [CI: -25, 55] compared to nnU-Net 7T. This resulted in the slope of the regression line increasing significantly from 0.68 for nnU-Net 7T to 0.90 for PLAn 7T (*P* < 0.05). This increased sensitivity corresponded to a moderate increase in mean lesion DSC (of 6%, *P* < 0.05), as well as a moderately improved expert lesion rating of 3.68 (CI: 3.33, 4.04) for PLAn 7T, compared to 3.53 (CI: 3.19, 3.86) for nnU-Net 7T (a 4% increase) with C-DEF-3T acting as reference.

**Figure 4.**
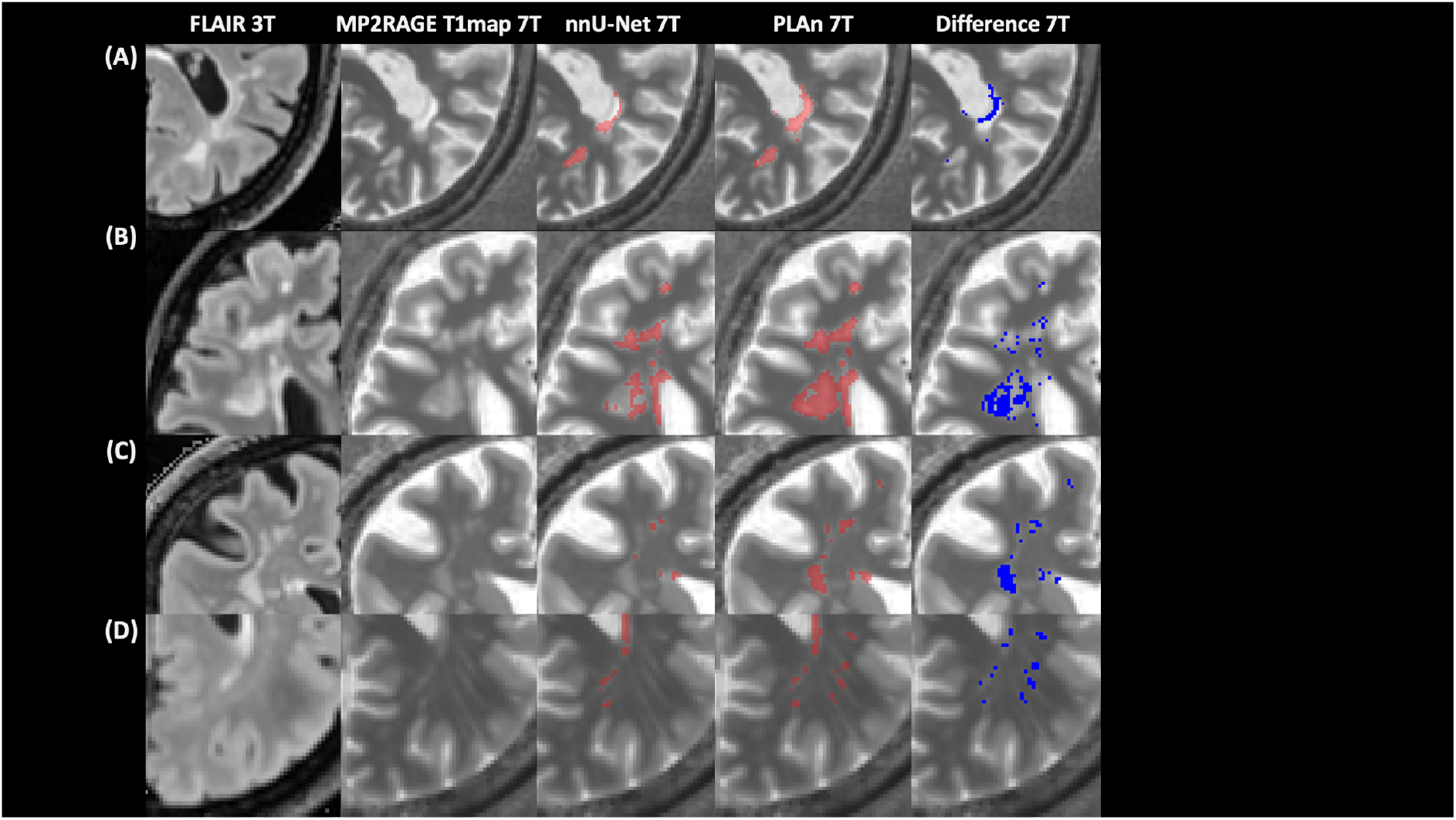
Improved lesion segmentation in PLAn 7T. Detailed comparison of segmentation results from nnUNET and PLAn methods from various participants, with 3T FLAIR and 7T MP2RAGE T_1_ map for comparison. Voxelwise subtraction of the lesion mask from nnUNet 7T and PLAn 7T methods with difference highlighted in red is overlaid on T_1_ map on the right.

## Discussion

This study demonstrates automatic whole-brain (including lesion) segmentation at 7T from a single, commonly gathered sequence without reliance on a priori assumptions or an atlas, which enables easy domain adaptation and implementation. In addition, this study presents the first effort to integrate skull stripping and whole-brain segmentation with the related but distinct task of lesion segmentation in 7T MRI. This contribution enables simpler and faster training, domain adaptation, and inference than previous methods in which these steps are handled by separate models or with the help of specialized sequences. These results show that nnU-Net, a publicly available medical image segmentation method, can produce good segmentations after training on only a handful of well-annotated training examples (*n* = 5), outperforming existing, methods for both skull stripping and tissue segmentation. Addition of a pre-training step to nnU-Net (in PLAn) significantly improved lesion segmentation performance by leveraging the readily available 3T pseudo-labels to mitigate limited 7T label availability. Importantly, a matching 3T dataset is only required for training, but not prospective application, of PLAn-7T.

Increased vulnerability to susceptibility artifacts, increased bias, and geometrical distortions can confound the coregistration of different 7T images which in-turn affect the modeling of local features across shared image space. Image distortions at air-tissue interfaces affects different acquisition schemes in different MRI sequences differently, which makes accurate whole brain registrations that is necessary for multi-contrast segmentation nearly impossible. Therefore, a segmentation method that minimizes these concerns by learning specific image features from scratch as well as by leveraging a single acquisition sequence, thereby requiring no coregistration step, was therefore advantageous to the multi-contrast segmentation method. Several sequences, such as multi-echo fast-spin or gradient-echo, might meet these criteria, nevertheless none are more useful than the robust imaging offered by MP2RAGE. The two inversion pulses can be tuned to optimize signal differences between tissue of interest (not only between GM and WM but also between lesion and CSF), and the postprocessing methods provide inherently coregistered T_1_w images. Finally, MP2RAGE sequence allows reconstruction of images at various simulated inversion times, which could offer further opportunities to differentiate other pathologies from normal tissue, but this not explored herein. For these reasons, MP2RAGE was chosen as the single sequence providing the multi-contrast input to the segmentation algorithm.

To benchmark PLAn against other methods, MRI scans of a cohort of study participants clinically diagnosed with, or suspected of having, MS were analyzed using several segmentation methods in parallel at both 3T and 7T. To ensure that the 3T and 7T segmentations were comparable, only study participants who had no new lesions detected between the two scans based on radiological reads were selected. Differences in lesion visualization between 3T and 7T, contrast attributes, and resolution dictated that a direct comparison between 3T and 7T would be imperfect.(10) This motivated the inclusion of the blinded neurologist ratings, which were done without a 3T reference in order not to skew the comparison of 7T methods against one another.

Both nnU-Net and PLAn produced excellent skull-stripping results at 7T, which is consistent with prior studies that have established the efficacy of U-Net-derived CNNs for brain extraction,(22, 23) including for patients with MS.(24) Both AFNI 7T and AFNI 3T generated results with similar limitations, including inclusion of some posterior orbital tissues and lack of inclusion of dorsal regions, with AFNI 7T producing more pronounced errors by over-stripping caudal regions such as the cerebellum and brainstem (Fig. 2D). Meanwhile, FreeSurfer 7T skull stripping significantly overestimated TIVs, while for 3 of the 20 inference-set participants, it included large regions of face and neck. These outliers were omitted from statistical analysis but indicate a clear shortcoming of this method.

Although some methods (FreeSurfer, C-DEF) enacted separate skull-stripping and segmentation steps, others (nnU-Net, PLAn) were designed to integrate these into a single step by treating non-brain voxels as a distinct segmentation class. While this did improve speed and convenience, skull-stripping discrepancies could not be easily separated from their downstream effects upon segmentation results. This effect was largely limited to CSF volumes, since this accounted for the vast majority of over- or under-stripped tissue. FreeSurfer 7T suffered from this effect most dramatically, although degradation of cortical structures in inferior brain regions also contributed to its overall error. It is likely that the observed degradation was due to inadequate mitigation of the strong 7T bias fields, since the observed degradation is systematic and localized to the temporal lobe and cerebellum regions in particular. Similar effects have been previously found in FreeSurfer analyses at ultra-high fields.(8) C-DEF 7T produced much better CSF segmentations but mostly failed to detect lesions, unlike its 3T counterpart. While these models both used MP2RAGE images, C-DEF 3T had the advantage of a FLAIR contrast, while C-DEF 7T did not. FLAIR contrast is invaluable for distinguishing lesions from GM and CSF when modeling tissue intensity signatures, as C-DEF does. As such, it appears that MP2RAGE images alone do not provide sufficient contrast for C-DEF to reliably separate lesions from other tissues. FLAIR images gathered at 7T are known to suffer from increased bias fields and artifacts, which along with any coregistration difficulties make them difficult to use for intensity-based segmentation tasks. Therefore, 7T FLAIR images were not collected or evaluated in this study. We did evaluate the efficacy of T_2_*-weighted 3D echo-planar imaging (EPI) scans as a potential independent contrast, but the images suffered from residual distortion errors near the sinuses, so they were not further considered.

In terms of segmentation methods, nnU-Net 7T clearly outperformed both FreeSurfer and C-DEF on all tissues at 7T, combining the advantages of both FreeSurfer (deep GM) and C-DEF (CSF, cortical boundaries) quite effectively. It also produced by far the most accurate lesion segmentations (Fig. 3). However, this comparison also revealed that nnU-Net, while more effective than the other baseline methods, still had substantial room for improved lesion detection. These shortcomings included a range of diverse lesion types, including diffuse WM T_2_-hyperintensities, periventricular lesions, and small punctate lesions throughout the brain.

In preliminary work, we investigated several strategies to boost nnU-Net 7T lesion performance (not explicitly presented here), including changes to data augmentation, training rate scheduling, and class sampling procedures, but these did not provide promising results. Another consideration was that while there were only a few (*n* = 5) manually edited 7T labels available for training, the nnU-Net method itself was developed on a set of public challenge datasets, which tend to be several orders of magnitude larger and substantially more diverse.(15) We therefore hypothesized that after pre-training the network on a larger, similar dataset with more abundant examples of lesion morphology, subsequent fine-tuning on our small, well-annotated 7T dataset could boost lesion performance and reduce overfitting (Supplementary Fig. 1). The C-DEF 3T segmentations were chosen as pseudo-labels for the pre-training step, since they were reasonably accurate as demonstrated previously (18, 25) and readily available, although not entirely free from all errors. In particular, they were derived partly from 3T FLAIR images, which prior studies have found can indicate more accurate lesion boundaries than those visible on 7T or 3T MP2RAGE scans (Fig. 4, leftmost column),(26) although the resulting volumes are typically highly correlated. It was therefore essential that the fine-tuning stage corrected these biases by learning from the 7T manually edited masks. To optimize the fine-tuning step, two parameters were evaluated: total training epochs and initial learning rate. We selected 125 epochs, which was the shortest training regime that remained stable, and an initial learning rate of 1 × 10^−4^, which was determined by cross-validation (Supplementary Fig. 2) to be the best compromise between preserving the pre-trained weights and learning new information from the 7T data. No significant advantages were observed by freezing any of the model layers, along the lines previously reported (27), and the final softmax layer was retrained. There is some evidence that for very small target data sets, enabling fine-tuning for the whole network may work best.(28)

The PLAn 7T model produced similar skull-stripping results to the default nnU-Net 7T, rather than those of AFNI 3T, which suggested that the fine-tuning was effective in correcting for errors in the 3T pseudo-labels. It also retained the other aspects of the 7T baseline model that were already optimal, including deep GM structures and cortical details. At the same time, the pre-training step was also effective in boosting lesion sensitivity without leading to the over-segmentation seen in the 3T baseline model (data not shown). While PLAn 7T did at times dilate existing lesion boundaries to cover the edges of diffuse WM hyperintensities, it also frequently added missing chunks of large diffuse lesions or detected lesions missed by all other methods, as shown in the qualitative analysis (Fig. 2, Fig. 4).

It was also observed that PLAn 7T displayed an increased tendency to denote very small T_1_ map-hyperintense features as lesions (Fig. 4C–D). While some of these features were determined to be blood vessels (Fig. 4D), many also appeared to be FLAIR-hyperintense lesions (Fig. 4C). While lesions smaller than 3 mm in diameter are typically excluded from diagnostic criteria for MS(29, 30) recent studies have found evidence that lesions below this threshold are more frequent in MS than in control subjects,(31) and they may even constitute the majority of lesions in MS.(32) These features may be of particular interest for high- and ultra-high field MRI, which enables higher resolution and signal-to-noise ratios, leading to higher lesion detection rates than at lower field strengths.(33) In any case, the occasional false positives caused by increased detection of perivascular spaces were dwarfed by the substantial gains in lesion detection elsewhere in the brain where previously missed by nnU-Net 7T. It should be noted that the final PLAn 7T lesion volumes remained lower than those of the C-DEF 3T segmentations, which was expected due to the inclusion of FLAIR scans at 3T.(26)

Several prior studies have employed CNNs derived from the U-Net framework to do cortical lesion segmentation(16) as well as whole brain segmentation without lesions(12) on 7T data, with encouraging results. However, this study is the first effort to achieve 7T whole-brain segmentation while including lesions, which are of particular interest as a marker for disease progression in MS (29) and other neurological diseases. (2, 3) Additionally, prior efforts toward lesion segmentation at 7T typically rely on either specialized T_2_*-weighted images(16) or a priori morphological rules(34), whereas PLAn 7T requires neither. By utilizing only images derived directly from a single sequence (MP2RAGE), PLAn 7T minimizes scan time and maximizes versatility while also avoiding the issue of coregistration error due to the distortion and artifacts that are particularly prevalent at 7T.

This study provides interesting insights regarding network training. For example, we demonstrated that nnU-Net can be trained from scratch with only a handful of labeled examples (*n* = 5), achieving reasonably robust performance superior to commonly used methods at 7T. This is a far smaller dataset than typically used for CNN training, even for MRI segmentation tasks, for which manually validated annotations are extremely time-consuming and expensive to generate. The proposed PLAn 7T approach could be considered an example of “few-shot” domain adaptation, in which only a few target domain examples are available. Our findings are consistent with prior studies, which indicate that transfer learning is often advantageous when the training dataset in the target domain is small.(27) One area for further investigation is whether a larger source or target domain training data set would significantly improve segmentation of lesions or other classes. A recent study of transfer learning for subcortical segmentation found that while just a few (1 to 3) images were sufficient for fine-tuning in most cases, smaller structures such as the amygdala and accumbens were most improved by additional data for fine-tuning.(35) Several recent studies have also used automated or semi-automated imperfect training labels (i.e., pseudo-labels) for model pre-training, followed by few-shot fine-tuning.(12, 36) These examples typically use larger source domain datasets than the one utilized in this study (*n =* 20), which may indicate that utilizing more pseudo-labeled data for pre-training could boost performance as well.

Prior studies have demonstrated the viability of transfer learning to mitigate scarce labels by leveraging data from other sites(12) or even artificially generated data.(37) However, the idea of leveraging data from lower field strengths to pre-train networks for 7T segmentation remains mostly unexplored. While one prior study has utilized a similar approach for cardiac cine MRI,(38) to our knowledge, this method has not previously been explored for brain MRI segmentation. This is somewhat surprising, since other types of domain adaptation, such as adaptation between different sites(36, 39) or contrasts,(28) have been considered. Another area for further exploration may be transfer learning across different neurological diseases. While this has been explored in a few prior studies,(28, 39) it is typically not decoupled from other domain adaptation tasks (e.g., changes in scanner or contrast), meaning it is yet underexplored.

One limitation of this study is that the methods evaluated were mostly assessed in an off-the-shelf fashion, with no hyperparameter tuning or modification unless explicitly stated. While dedicated fine-tuning of any of these methods may yield improved results for a specific cohort, such extensive optimization is beyond the scope of this study. In addition, this comparison was limited to a single cohort of MS study participants with both 3T and 7T MRI scans. This was done to allow for detailed comparison as well as a reliable reference point (C-DEF 3T) without large-scale manual annotation, which can be incredibly expensive and tedious. These labels could not be treated as a true 7T gold standard owing to the inherent differences between segmentation at 3T and 7T, as well as with and without FLAIR contrast. Future work will address these limitations by extending such a comparison on multi-site data including other disease cohorts and refining the method for pseudo-label and gold standard generation. Initial results are promising (data not shown), but it is possible that images from other manufacturers and large sequence parameter changes will require retraining of the algorithm.

In this study, a state-of-the-art deep learning method, was developed and applied to MP2RAGE 7T MR images with the goal of obtaining fast and reliable whole brain segmentations. This method enables skull stripping and segmentation in a single inference step and is better able to detect MS lesions than existing methods for 7T MRI segmentation. By employing only multi-contrast techniques such as MP2RAGE, it is readily scalable and adaptable to use in a wide variety of conditions and performs robustly against any registration errors. Performance of 7T segmentation was boosted and training label scarcity for lesion segmentation overcome by incorporating a pre-training step using results from a robust 3T segmentation as pseudolabels before finetuning a transfer-learning method. We present these findings as a blueprint for acquisition of fast, accurate, and useful volumetric markers from 7T MRI data for use in clinical and research settings.

## Data Availability

The data and code used here may be obtained with Material Transfer Agreements, subject to NIH policy on data sharing.

## Acknowledgments

This research was supported by the Intramural Research Program of the National Institute of Neurological Disorders and Stroke.

This work utilized the computational resources of the NIH high-performance computing (HPC) Biowulf cluster (http://hpc.nih.gov). We thank the NINDS Neuroimmunology Clinic for recruitment and evaluation of study participants and the NIMH Functional MRI Facility for assistance with MRI data acquisition. The authors report no competing interests directly related to this study.

---

**Supplementary Figure 1:**
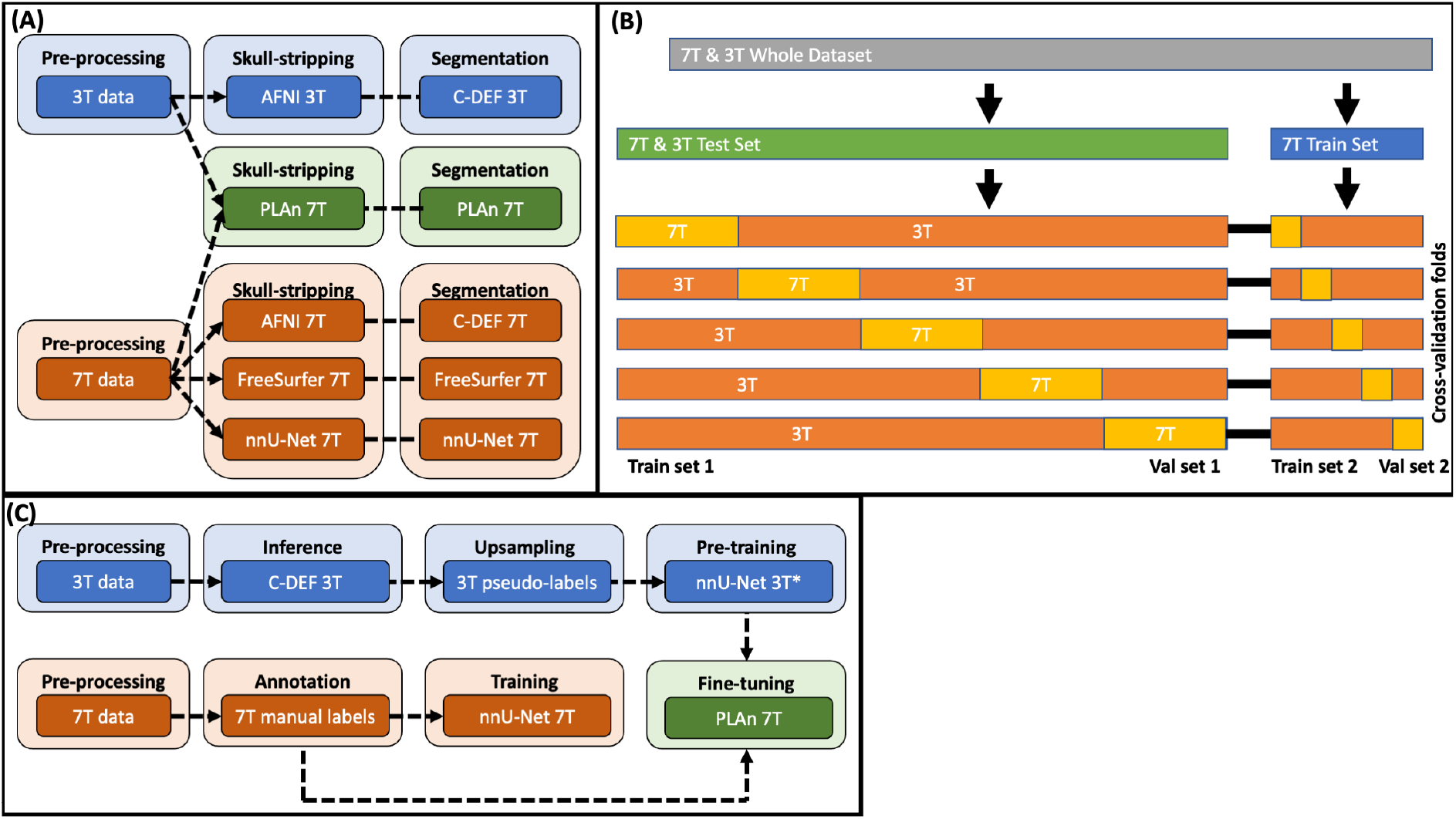
Methodology workflow. **(A)** skull stripping and segmentation comparison for various methods, **(B)** dataset splits used for PLAn 7T, and **(C)** transfer learning scheme used for PLAn 7T.

**Supplementary Figure 2:**
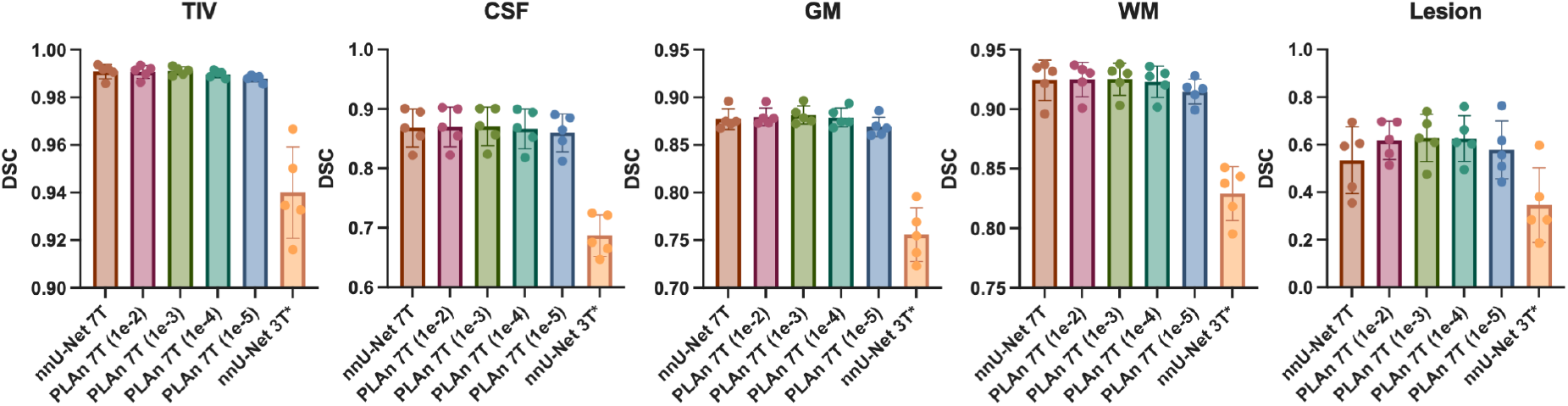
Cross-validation Dice similarity coefficient (DSC) plots for PLAn 7T fine-tuning learning rate optimization. Learning rate for PLAn fine-tuning indicated in parenthesis. nnU-Net 3T* denotes the network after pre-training with only pseudolabels (i.e., no fine-tuning).

